# Differentials in clinical severity and other patient activity indicators amongst Black and South Asian cancer patients in England

**DOI:** 10.1101/2022.06.24.22276862

**Authors:** Steffan Willis, Pedro Figueiredo Aparicio, Rhoda Steel, Gaetan Leblay

## Abstract

The link between ethnicity, deprivation and health inequalities is well-established. The relationship between ethnicity and cancer is more complex and influenced by a variety of socio-economic, cultural and physiological factors. Understanding the relationship between ethnicity and patient care indicators for specific cancer types is vital if NHS England is to meet the UK government’s stated priority to reduce health inequalities as it recovers from COVID-19.

This paper explores the impact of ethnicity on clinical severity, treatment costs and a range of patient activity indicators across three cancer types – chronic lymphocytic leukaemia, multiple myeloma and prostate cancer.

The paper uses a dataset derived from the Hospital Episodes Statistics (secondary care) database covering 2016/17 to 2020/21,. This enabled the differential impact of the pandemic on ethnic minority patients to be considered. The data was aggregated by ethnicity and deprivation quintile at a national and Integrated Care System (ICS) level. Clinical severity was proxied using co-morbidity and complications (CC) scores. Multivariate linear regression (OLS) models were used to explore the associations with ethnicity.

Black and South Asian patients CC scores were 12.2% and 15.8% higher than the population average (4.1). Controlling for socio-economic deprivation, South Asian patients had higher average clinical severity (+0.57, p<0.01). In addition, ICSs with large South Asian populations were associated with higher CC scores (+0.69, p<0.01). Treatment costs were higher for Black prostate cancer patients with interventions (+£842, p<0.001) and South Asian multiple myeloma patients (+£1686, p<0.001). Both Black and South Asian patients tend to have more spells in hospital. COVID-19 saw total inpatient admissions fall by 18.9%. Black and South Asian inpatient admissions fell by 1.9 and 2.9 percentage points more than the national average respectively. Average clinical severity increased by 7.1% with the largest increase amongst South Asian (+11.5%) and Black (+8.1%) patients.

The higher clinical severity in South Asian patients and higher treatment costs in Black patients observed in this study are not accompanied by significant variations in patient activity indicators, which may point to drivers associated with delays to diagnosis or barriers to access to primary care.

## Introduction

### Ethnicity and Health Inequalities

There is a well-established literature linking ethnicity to a range of health inequalities in England [1]. The relationship between ethnicity and ill-health is complex and mixed. Higher rates of socio-economic deprivation, amongst ethnic minority groups contribute to higher levels of ill-health [2][3][4]. However, there are mitigating factors such as lower rates of smoking and alcohol consumption, especially amongst first-generation migrants. Physiological differences also play a role in the incidence of specific conditions, including several cancer types.

Ethnic minorities are more likely to report poor health and limiting long-term illness than White British people, with rates especially high amongst the South Asian population. A significant proportion of the variation in self-reported ill-health is explained by higher levels of socio-economic deprivation amongst ethnic minority communities [5]. Table 1 shows the percentage of people living in the 10 percent most deprived neighbourhoods in England split by ethnicity and type of deprivation [2]. Black and South Asian people are significantly more likely to live in the most deprived areas – 31.1 percent of the Pakistani population live in the most deprived decile. Although the health deprivation and disability domain shows some of the lowest rate differentials, this is associated with the low median age of neighbourhoods with large black and South Asian populations. Controlling for socio-economic status, elderly individuals of Pakistani and Bangladeshi background have significantly higher odds of reporting health issues which limit typical activity than the White British population [6].

**Table 1:**
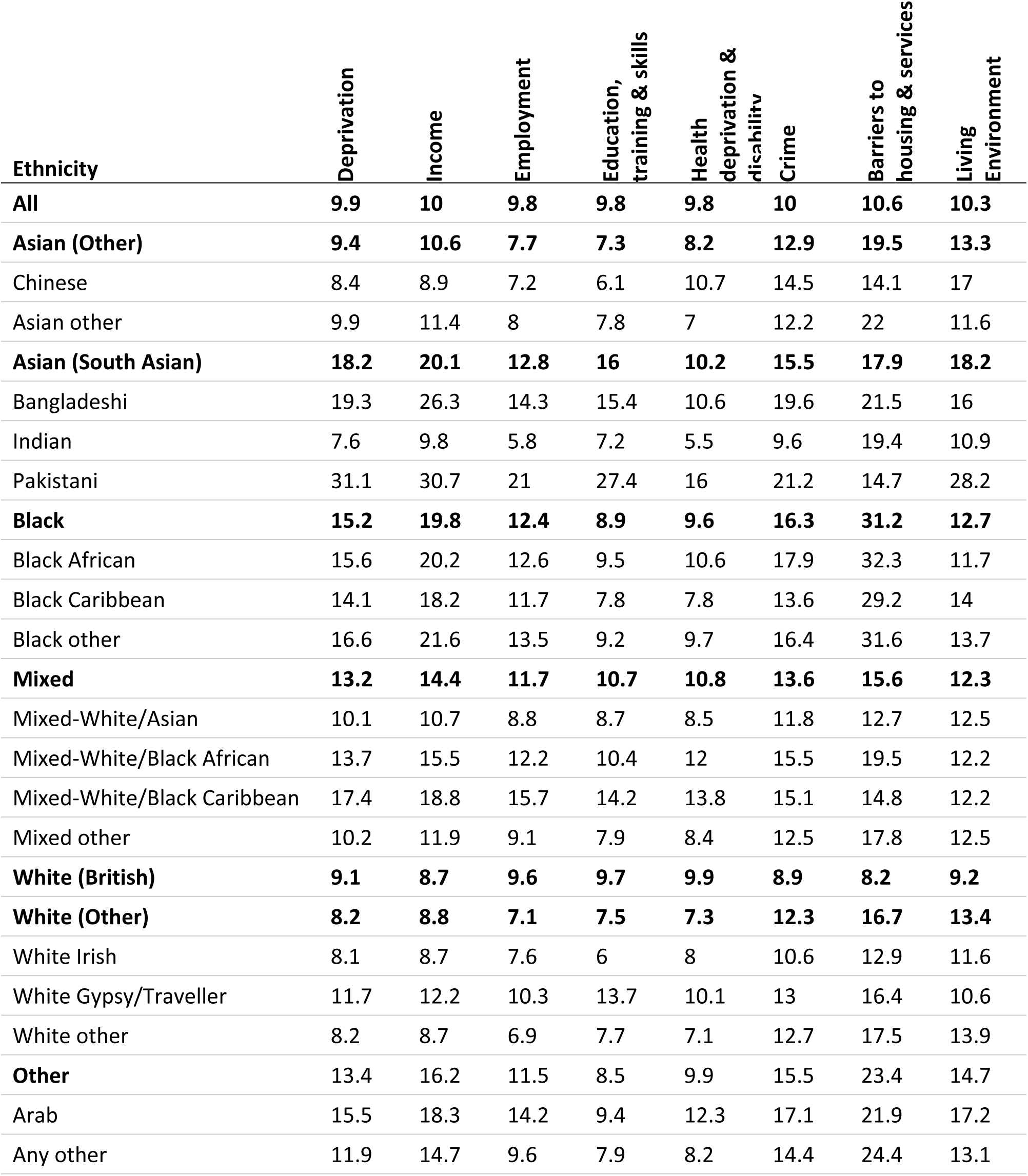
Percentage of people living in the most deprived 10 percent of neighbourhoods by ethnicity and deprivation type (Index of Multiple Deprivation)

Health inequalities are more severe in London than in the rest of England reflecting higher levels of socio-economic inequality. Rates of long-term limiting illness amongst Bangladeshi women are 30 percent higher than White women in London, compared with 15 percent higher in the rest of England [5] . Ethnic minority populations tend to be concentrated in major urban areas. Table 2 shows the top 10 Integrated Care System (ICS) areas by Black, South Asian and White population – 58.7 percent of the Black population live in the five ICS areas that cover Greater London [7]. Ethnic minorities make up 65 percent of the population in the North West London ICS, but less than 10 percent in almost two-fifths of ICSs. Policies aimed at addressing ethnic health inequalities need to meet the specific needs of local ethnic minority communities.

**Table 2:**
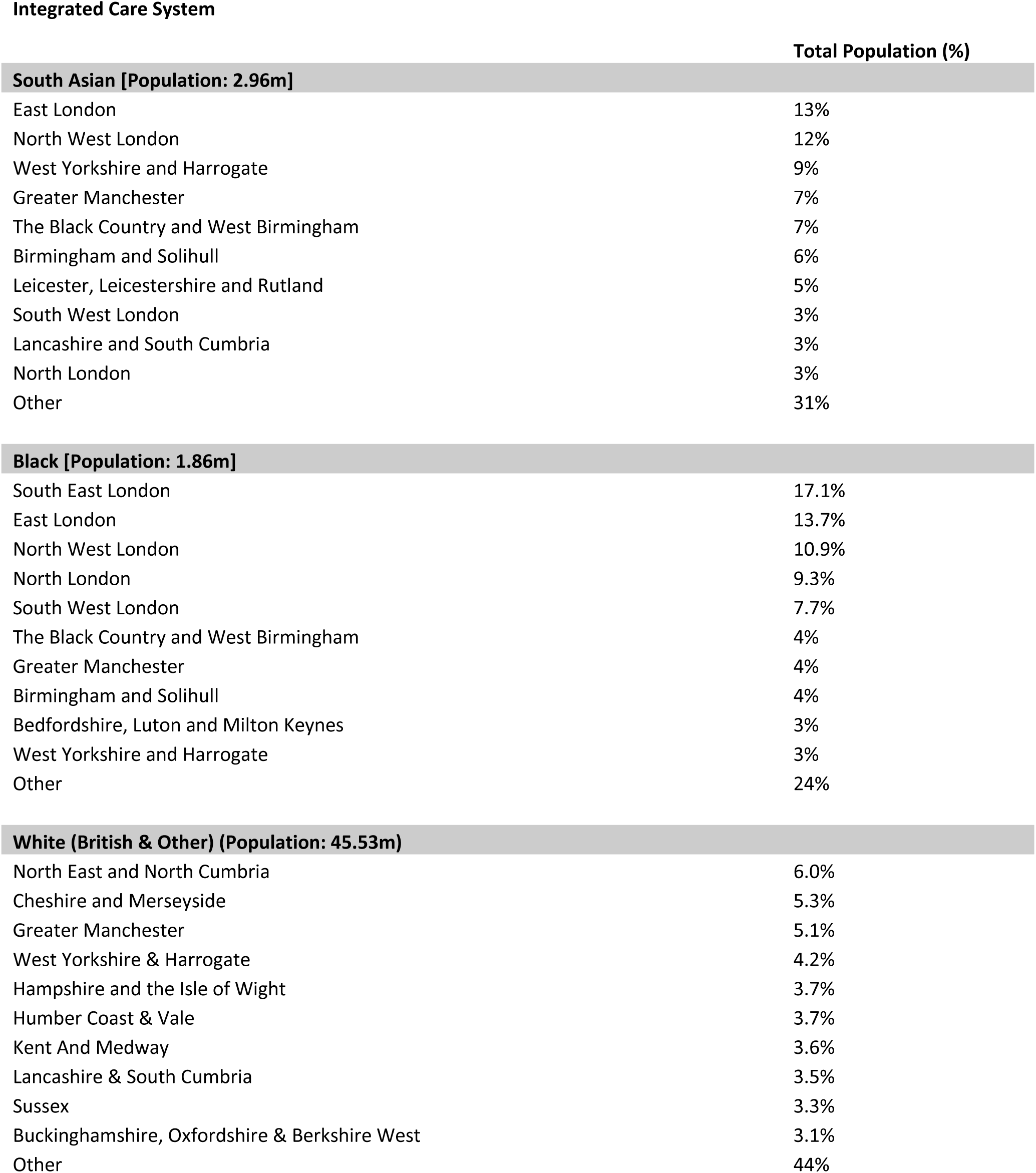
Top 10 Integrated Care Systems of major ethnic groups in England showing percentage of total population.

Equity in access to care is a foundational principle of the NHS. In contrast to many other health systems care is provided free at the point of use. However, paradoxically, the lack of economic barriers to accessing healthcare in the UK can at times lead to complacency when it comes to addressing broader socio-cultural factors which limit access to healthcare [8]. Policies aimed at addressing ethnic health inequalities in England have a long history, and are supported by strong legal frameworks. The Equalities Act of 2010 placed a legal responsibility on the NHS to advance equality in health and social care across different ethnic groups. However, despite these policy commitments evidence of health inequalities remains. A review of healthcare commissioning stakeholders between 2010 and 2012 found that the national policy agenda was frequently marginalised in favour of other commissioning priorities and treated with ambivalence by NHS leaders. Local commissioning groups lacked high-quality data on ethnic differences in patient care and outcomes. Many stakeholders were unclear on the specific actions required to address ethnic inequalities as distinct issues separate from the challenges associated with socio-economic deprivation [8].

COVID-19 acted as a stark reminder of the ongoing presence of ethnic health inequalities in the UK. Many of the factors which contribute to greater ill health in ethnic minority communities are also linked to increased risk of infection and higher mortality rates. There is significant evidence that during the early stages of the COVID-19 pandemic ethnic minorities, particularly Black and South Asian individuals, suffered disproportionately high incidence and mortality rates [9][10][11]. Between June 2020 and May 2021, the age-adjusted mortality rate for the Asian population in England was 2.1 times higher than the White population, and 1.6 times higher for the Black population [12]. Dispiritingly, these patterns mirrored those observed, at a much smaller scale, during the 2009 pandemic influenza flu season [13]. In addition, there is evidence that the ethnic minorities suffered greater economic and social costs as a result of COVID-19 restrictions [14].

As the debate shifts from dealing with the direct impact of COVID-19 to the long-term recovery of the health care system, ensuring that specific ethnic health inequalities are fully understood and given policy priority is essential if the NHS is to deliver on its founding principles. In July 2021, the UK government published its Life Sciences Vision, outlining its policy priorities for the next decade [15]. Addressing health inequalities, particularly through the use of NHS data partnerships, was listed as a key objective. To this end, NHS England and the NHS Confederation launched the NHS Race and Health Observatory in May 2020 to investigate the impact of race and ethnicity on people’s health and to identify and tackle specific health challenges facing ethnic minority communities [16]. The chair of this organisation identified the digital data analysis of health inequalities across all levels of the NHS as “fundamental” to this mission [17]. This paper seeks to provide a timely contribution to the debate through a quantitative analysis of the relationship between ethnicity and patient care outcomes in three cancer types – chronic lymphocytic leukaemia, multiple myeloma and prostate cancer.

### Ethnicity and Cancer

The relationship between ethnicity and cancer is complex with a variety of competing socio-economic and physiological factors at play. The combined age-standardised cancer incidence rates are higher amongst the White population than other ethnic minority groups. The exception to this is Black men who have rates roughly equivalent to those of White men, although this is largely driven by significantly higher incidence of prostate cancer. Cancer incidence (all forms) amongst Black and Asian women is around 40 percent lower than for White women [18]. That said, there are significant differences in incidence, mortality and age of onset across different cancer types [19][20][21].

#### Chronic Lymphocytic Leukaemia

Chronic lymphocytic leukaemia (CLL) accounts for around 1 percent of new cancer diagnoses in the UK [19]. Black and South Asian populations tend to have lower incidence rates than the White population [22]. Data from the US Surveillance, Epidemiology and End Results Program (SEER) suggests that the average life time risk of being diagnosed with CLL is 1.9 times for White Americans than Black American and 3.7 times higher than Asian Americans [23]. The five-year survival rate for patients in England with CLL is 66.5 percent for males and 72.5 percent for females[19]. Survival and mortality rates split by ethnicity are not readily available.

#### Multiple Myeloma

Multiple myeloma is the 19^th^ most common form of cancer in the UK accounting for 1.6 percent of cancer diagnoses and 1.9 percent of deaths. Incidence rates increase with age and are higher in males than females, 73 percent of diagnoses are in those aged over 65 [20]. Incidence rates for people from White and Asian ethnic backgrounds are broadly comparable. However, incidence rates in the black population are 2-3 times higher. This pattern is also observed in the US, Black Americans have a lifetime risk of diagnosis 1.8 times higher than White Americans [23]. This data also suggests that Asian Americans have a lower risk of being diagnosed with multiple myeloma, however, caution is required in translating this to a British context given the different composition of the Asian population in the two countries. Individuals of South Asian descent make up a far higher share of the Asian population in the UK (71 percent) than the US (21 percent). Black Americans have an earlier average age of onset, but also higher survival rates [24]. The five-year survival rate for patients with multiple myeloma is 52.3 percent, falling to 29.1 percent after 10-years. Survival rates fall as age increases and are marginally lower for males than females [20].

#### Prostate Cancer

Prostate cancer is the most common cancer in the UK male population. It accounts for 27 percent of male cancer diagnoses and 13.3 percent of deaths. Incidence rates are extremely low in younger age groups but rise rapidly with age from the mid-40s. Three-quarters (75.6 percent) of diagnoses are in those aged over 65 [21]. Prostate cancer is much more common amongst Black males than other ethnic groups with a lifetime risk of being diagnosed of 29.3 percent [30]. This compares to 13.3 percent for white men and 7.9 percent for Asian men. Data from the US SEER Program suggests that Black men have an earlier average age of onset [23]. The five-year survival rate for patients with prostate cancer is 86.6 percent, falling to 77.6 percent after 10-years. Survival rates decline with age [20].

#### Socio-Economic Deprivation

Increased levels of socio-economic deprivation are associated with lower diagnosis rates, particularly for male patients. The most deprived quintile has an age-standardised incidence rate 17.1 percent lower than the least deprived quintile. If all areas had incidence rates equivalent to the least deprived quintile we would expect to see and additional 3,079 cases of prostate cancer diagnosed each year [26]. This does not reflect lower actual incidence amongst more deprived groups, rather it is likely the result of variation in access to and utilisation of heath services and prostate cancer screening [27].

#### Experience of Care

Ethnic minority patients in England report lower satisfaction rates and more negative experiences of cancer care. Controlling for socio-economic deprivation, Black and Asian patients were less than half as likely as White British patients to rate their care as ‘Excellent’. They also had lower levels of confidence and trust in medical staff and were less likely to fully understand medical advice [28] Cancer patient experience is worse in London than in the rest of England, although this was not significant impacted by a patient’s ethnicity [29]. Summary analysis of more recent National Cancer Patient Experience Survey data suggests that these issues have been largely unaddressed [30].

Different aspects of the relationship between ethnicity and cancer have been explored in a large, but still incomplete literature. At the population-level, a relatively large body of work exists documenting the relationship between ethnicity, deprivation and cancer incidence and mortality across a variety of cancer types, although many of these studies are limited by the size of ethnic minority patient populations. This is supplemented with a small but robust literature exploring the socio-cultural barriers such as language, cultural stigma and knowledge of symptoms which may impact help-seeking and diagnosis amongst ethnic minority groups. These tend to be focused on the primary care sector as the initial interface with healthcare services. Analysis of the secondary care sector has largely focused on patients experience of cancer care drawing upon self-reported survey data. However, whilst important, this often fails to capture the more direct links between ethnicity and a variety of patient activity indicators and outcome measures. This paper seeks to address some of these gaps by examining the relationship between ethnicity and average clinical severity, treatment costs and a variety of patient activity indicators for three specific forms of cancer - chronic lymphocytic leukaemia, multiple myeloma and prostate cancer.

## Methods

### Data Source

The primary data used in this paper is drawn from the Hospital Episode Statistics (HES) database. The HES database contains details of all in- and outpatients admissions and appointments in NHS hospitals in England including detailed clinical, patient, administrative and geographical information. The dataset used in this report covers the 5 fiscal years 2016/17 to 2020/21 and contains information on the hospital activity of all patients with a relevant ICD-10 diagnosis code (see Table 4) for whom a comorbidity and complications (CC) score could be extracted. The data was augmented with deprivation data from the English Index of Multiple Deprivation mapped to the patient’s residence using Lower Layer Super Output Areas (LSOAs). Data querying, aggregation and suppression was conducted by a private provider, Wilmington Healthcare, under licence by NHS Digital.

**Table 3:**
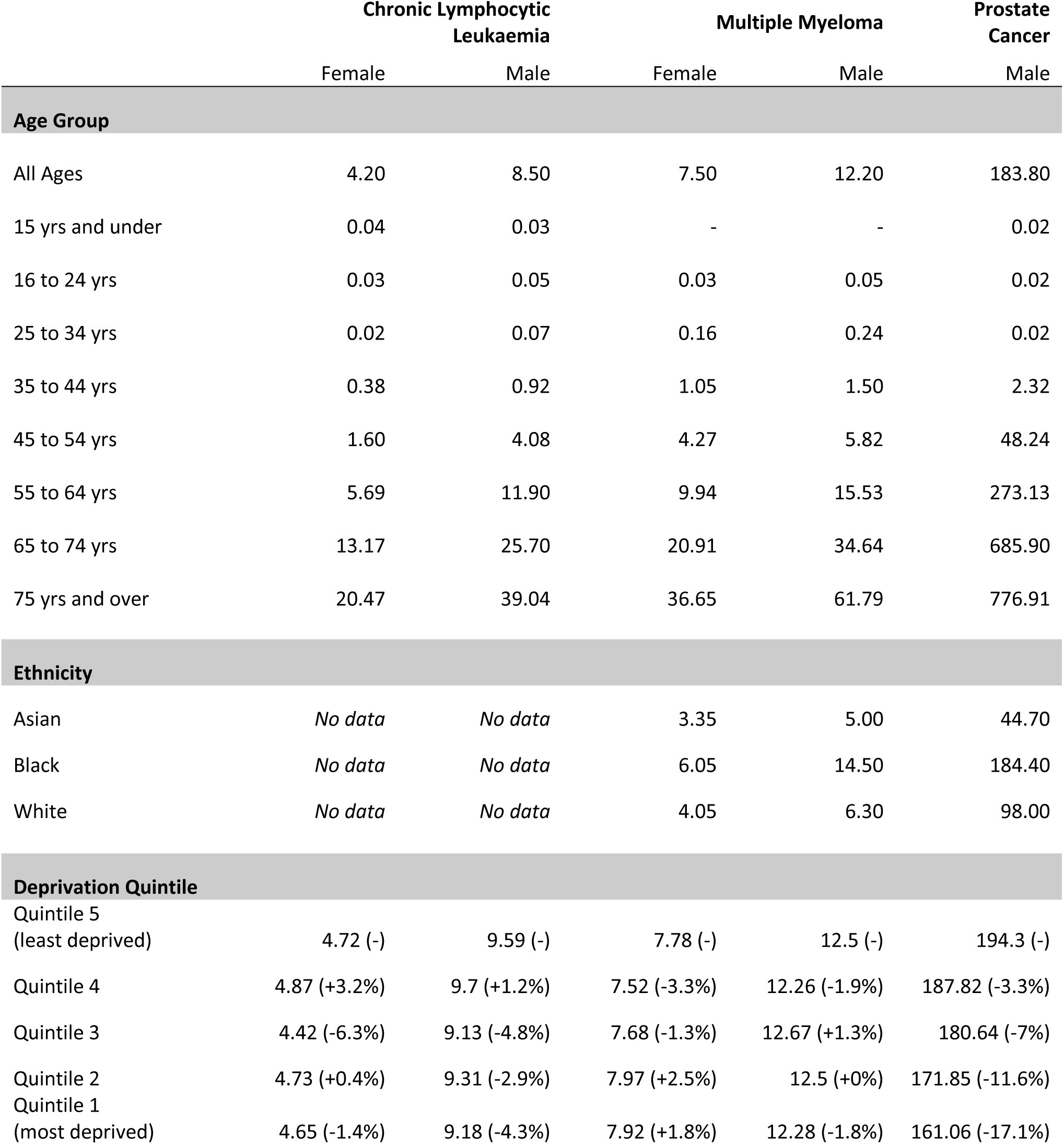
Age-standardised incidence rates per 100,000.

**Table 4:**
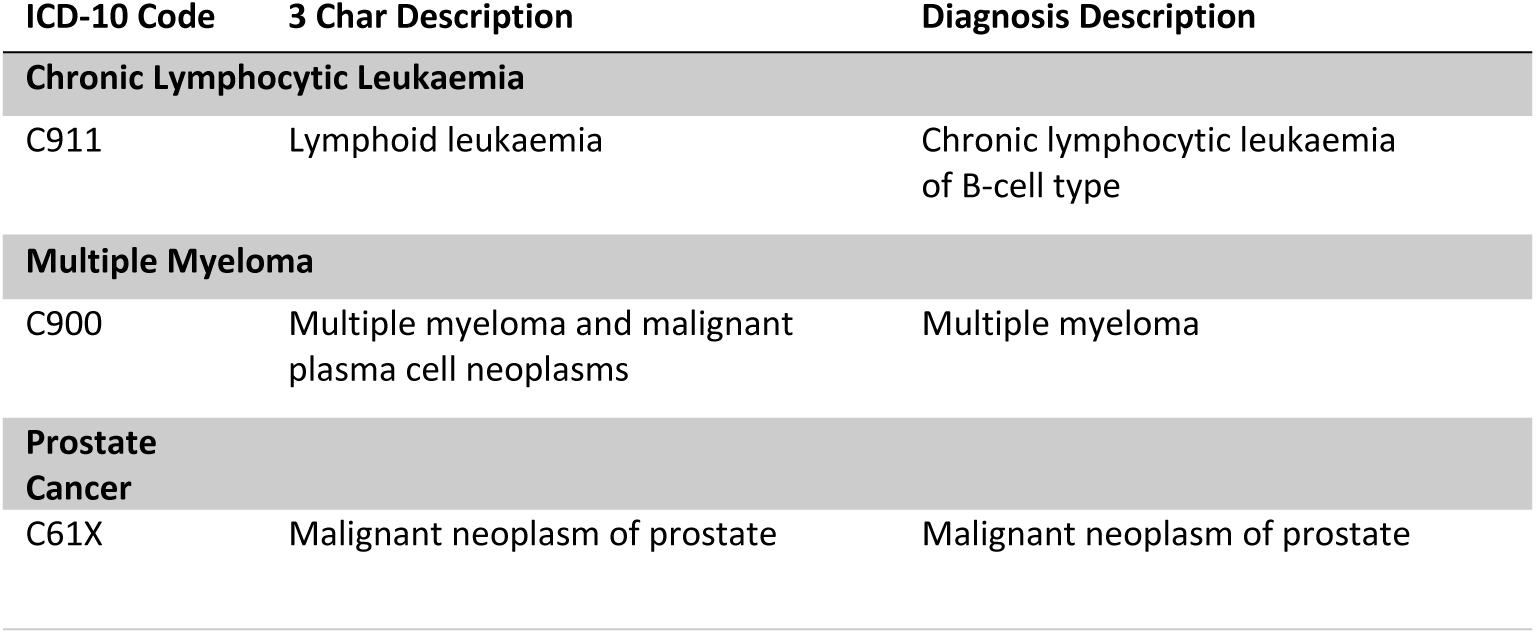
Relevant diagnoses and health resource groups.

| ICD-10 Code | 3 Char Description | Diagnosis Description |
| --- | --- | --- |
| **Chronic Lymphocytic Leukaemia** |  |  |
| C911 | Lymphoid leukaemia | Chronic lymphocytic leukaemia of B-cell type |
| **Multiple Myeloma** |  |  |
| C900 | Multiple myeloma and malignant plasma cell neoplasms | Multiple myeloma |
| **Prostate Cancer** |  |  |
| C61X | Malignant neoplasm of prostate | Malignant neoplasm of prostate |

Patient activity measures were aggregated by ethnicity and deprivation quintile at a national and ICS level for each of the fiscal years and for the combined five-year period. To protect the anonymity of patients suppression protocols were applied to the aggregated data. The choice of ICS areas as a sub-national geographic area to be included in the analysis was driven by the need to balance greater geographic granularity against smaller patient populations and the associated increase in data suppression. There are 42 ICS areas in England with populations of between 1 and 3 million people. ICSs are a key aspect of the NHS Long term Plan [31]. Values between 1-7 for the number of patients and spells in hospital were suppressed, values greater than 7 were been rounded to the nearest 5. Per patient measures were rounded to the nearest whole number or the nearest five in the case of average cost per patient. In addition, in order to assess the variation in clinical severity across demographic groups comorbidity and complications (CC) scores were extracted from the Health Resource Group (HRGs) bands associated with each patient admission. CC scores are specific to HRGs and denote the level of comorbidity and complication associated with the admission. Both CLL and multiple myeloma have a one to one relationship with a HRG whereas prostate cancer is split across two HRGs according to whether the patient required interventions.

The dataset covers three distinct cancer types – chronic lymphocytic leukaemia, multiple myeloma, and prostate cancer. The dataset (see Table 5) contains aggregated data from 59,830 unique patients with the distribution by ethnicity broadly in line with expectations given the epidemiological profiles of these cancer types and the demographic profiles of different ethnic groups in the UK. Ethnic minorities make up 23 percent of those aged under 65, but only 8 percent of those aged over 65 [32].

**Table 5:**
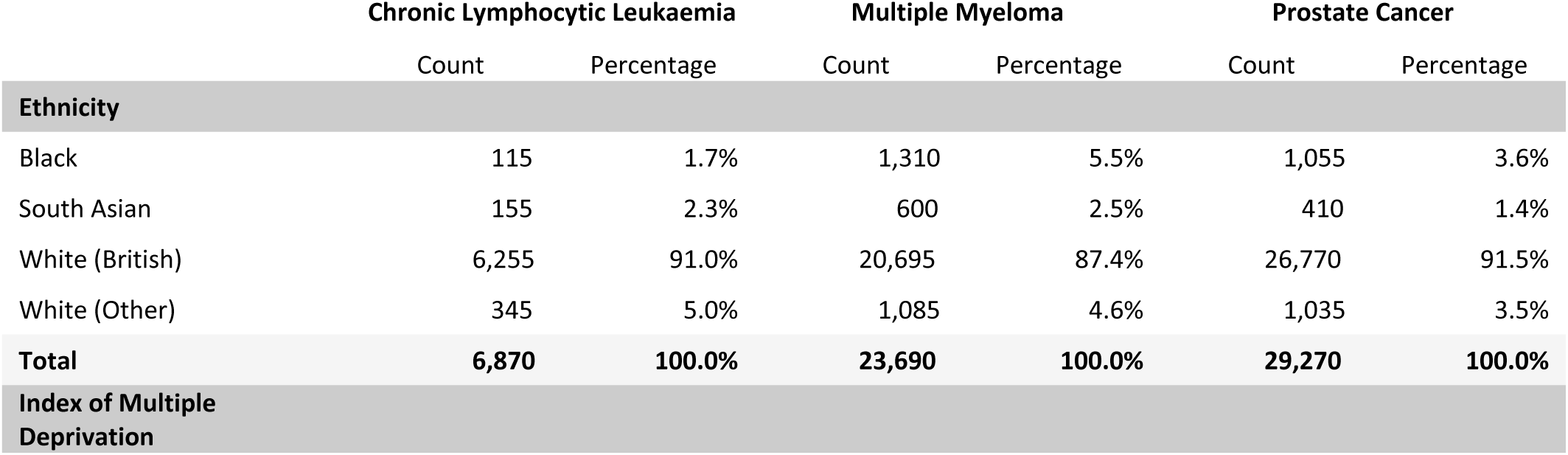
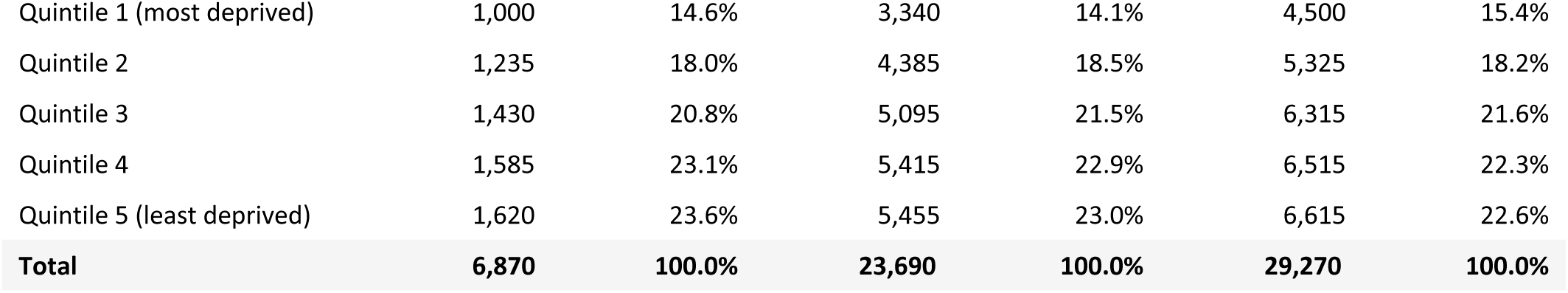
Patient population split by Ethnicity and Deprivation Quintile.

### Measures

#### Dependent variables

In order to develop a holistic view of the impact of ethnicity on secondary cancer care and outcomes a number of different dependent variables were considered. The primary focus of the analysis is the impact of ethnicity on *clinical severity*. Clinical severity is estimated using average CC score as a proxy. Due to different grading systems across HRGs, CC scores have been normalised on 1-10 scale using a max-min normalisation process. The differential between this score and the population average was then calculated, allowing different HRGs to be compared directly. *Treatment costs per patient* are closely linked to CC scores due to their role in the NHS’s payments framework. However, they provide a healthcare system focused, rather than patient-focused, measure enabling the relative allocation of resources to be assessed. Three in-patient activity indicators are considered - *spells per patient (in-patient hospital admissions), average wait duration, and mean length of stay*.

#### Independent Variables

At a national level two independent variables are used – ethnicity and deprivation. *Ethnicity* is the primary variable of concern. The analysis is focused on the four largest ethnic groups in England – White British, White (Other), South Asian and Black. As a result of the relatively small patient populations the ‘Asian (Other)’, ‘Mixed’ and ‘Other Ethnic Group’ categories were excluded from the analysis. These accounted for 1.3 percent of the CLL patient population, 3.4 percent for multiple myeloma and 2.2 percent for prostate cancer. In addition, patients categorised as ‘Unknown ethnic group’ were removed, these represented between 5.2 percent and 7.4 percent of the patient populations.

The White (Other) ethnic grouping is the largest ethnic minority group in the UK with an estimated population of 4.15 million in 2019 [32][33]. However, this group contains significant levels of demographic and socio-economic variation including both a large established White Irish population who have similar health outcomes to the White British majority [6] and more recent migrants from Eastern Europe who tend to be younger with lower socio-economic resources. For the purposes of descriptive analysis the White Other and White British categories are presented independently, however, for these ethnic groups were combined when constructing the multivariate linear regression models. In these models, the ethnicity variable was recoded into binary variables representing the South Asian and Black ethnic groups with the ‘White’ binary variable excluded to avoid issues of multi-collinearity.

*The Index of Multiple Deprivation (IMD)* quintile is a measure of deprivation based on a variety of socio-economic and environmental factors. It includes weighted measures of income, employment and economic inequality, access to housing and services, and health inequalities and is calculated at a local-level in areas with populations of approximately 1500 people. Every area is assigned a score and rank. The Index of Multiple Deprivation are national statistics published by the Ministry of Housing, Communities and Local Government, the most recent publication of the indices was in 2019 [34]. Patient data has been aggregated based on the quintile rank of the area in which they live, with the lowest quintile covering the 20 percent most deprived areas in England. The variable has been recoded into a binary variable, with the lowest two deprivation quintiles representing ‘high’ deprivation areas.

Due to issues of patient confidentiality and data suppression, the data could not be cross-tabulated using factors such as age or gender, this is a significant limitation of the dataset. To partially address this gap, the ICS-level models include the *percentage of the ICS population aged over 65* as an independent variable.

This variable also acts as measure of the White British population due to the higher correlation between these variables (R^2^ = 0.77). In addition, to explore the effect of single-ethnicity concentration, the ICS- model for clinical severity includes the *South Asian and Black population percentages.* Demographic data was sourced from NOMIS drawing on the 2011 Census [7]. Both ethnic groups are highly concentrated major urban areas (see Table 2) – over half the Black population live in London. Each of these variables was normalised on a 0 – 1 scale using max-min normalisation. This was done to improve the comprehension of the regression coefficients. Finally, when considering treatment costs, a *‘London’* binary variable which captures whether an ICS-area is in London was included in the model due to the higher costs of delivering health services in the capital.

For the descriptive analysis, cross-tabulations were used to analyse the distribution of average clinical severity across different ethnic groups. To explore the association between ethnicity and the dependent variables ordinary least-squares (OLS) multivariate linear regression models were used to estimate the strength of the relationships and explanatory power. All models estimated using the scikit-learn module in Python [35][36].

## Results

### Patterns of clinical severity across the population

Table 6 shows the average normalised CC scores for each of the major ethnic groups and the differential between that score and the population average. A positive differential reflects higher average clinical severity within that population grouping. The population average is closely aligned to average scores of the White British population.

**Table 6:**
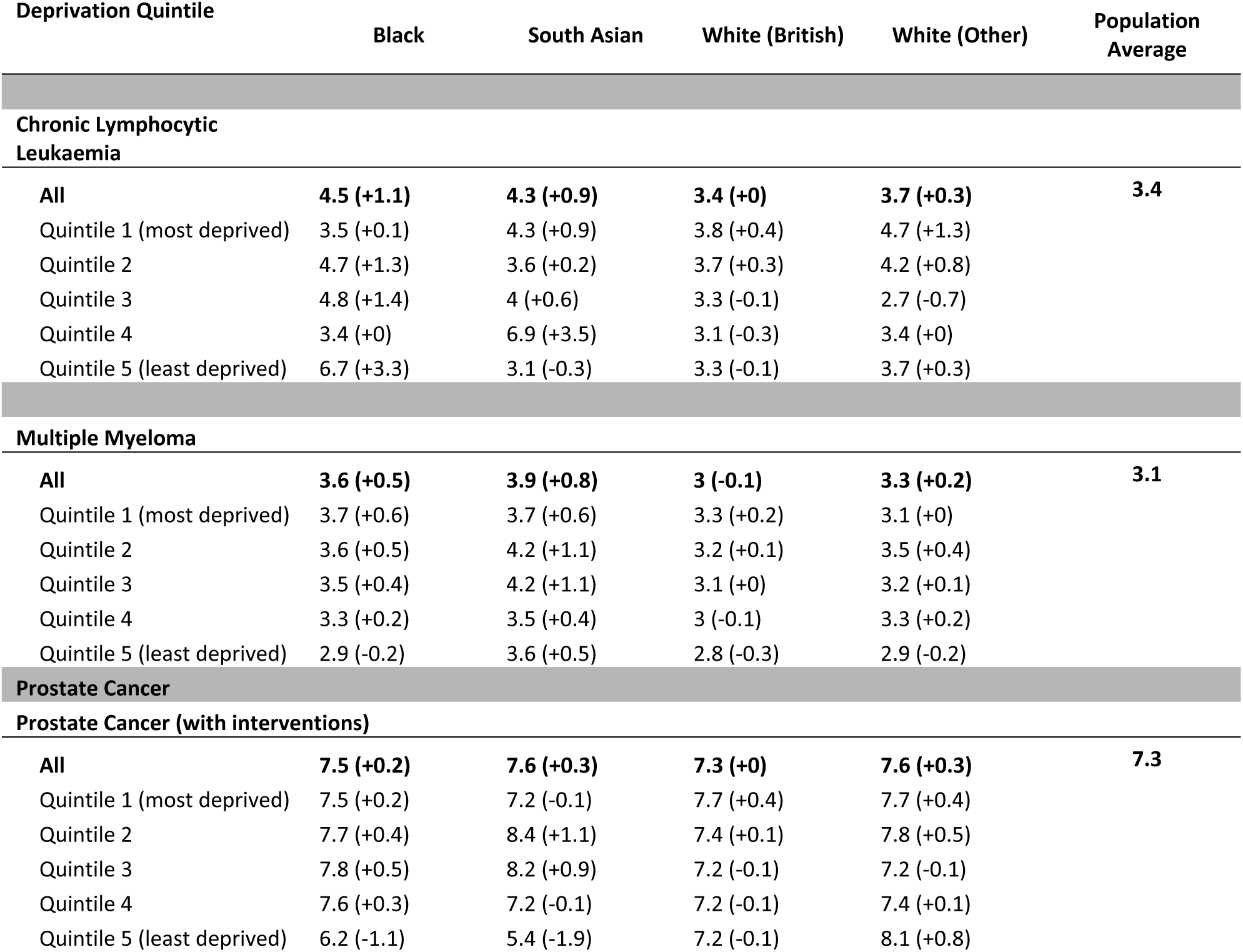
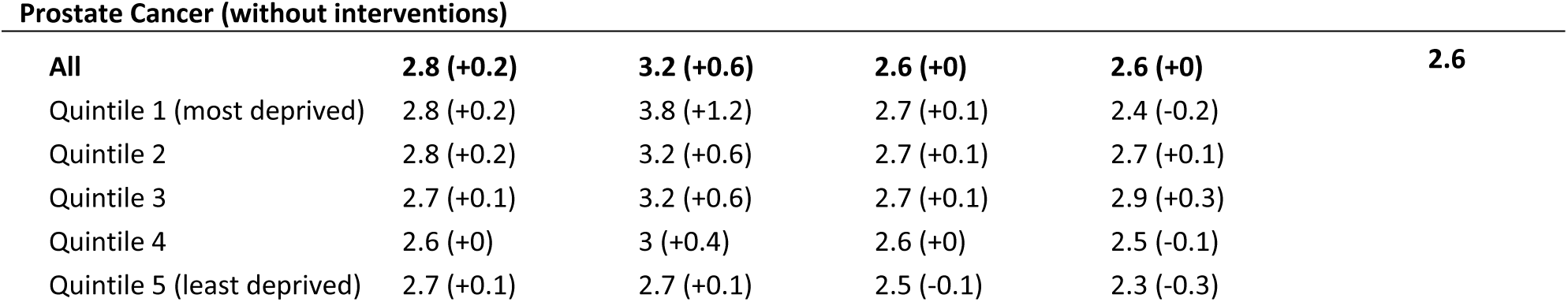
Average normalised complications and co-morbidity score (clinical severity) and differential with population average score by major ethnic group and deprivation quintile.

All ethnic minority groups have average CC scores equal or higher than the population average for all disease areas. The South Asian ethnic group has the highest average clinical severity for all disease areas with the exception of chronic lymphocytic leukaemia (CLL). Across all disease areas average CC scores were 20 percent higher than the population average. There was more variation in results for the Black population with relatively high scores for both chronic lymphocytic leukaemia and multiple myeloma but a smaller differential amongst prostate cancer patients, average CC scores were 11.8 percent above the population average. The White (Other) population displayed consistently higher clinical severity than the population average, however, the size of effect was relatively small (+6.8 percent). Higher levels of socio-economic deprivation are generally associated with higher clinical severity.

Table 7A present the results of a statistical model used to examine the relationship between ethnicity and clinical severity at a national level for all previous cancer types. The model controls for higher clinical severity in areas with higher deprivation. When data from all patients is are combined, being of South Asian ethnicity was associated an average CC score differential of +0.58 percentage points higher than population average with the result statistically significant at a 99% confidence level (CL). Being Black and living in a high deprivation area were both associated with higher clinical severity (p<0.1) although the magnitude of the effect was roughly half as strong as that of the South Asian variable. The model had an R^2^ of 0.147 which given the variation across different diagnosis groups and the issues associated with aggregation and suppression is to be expected.

**Table 7A:**
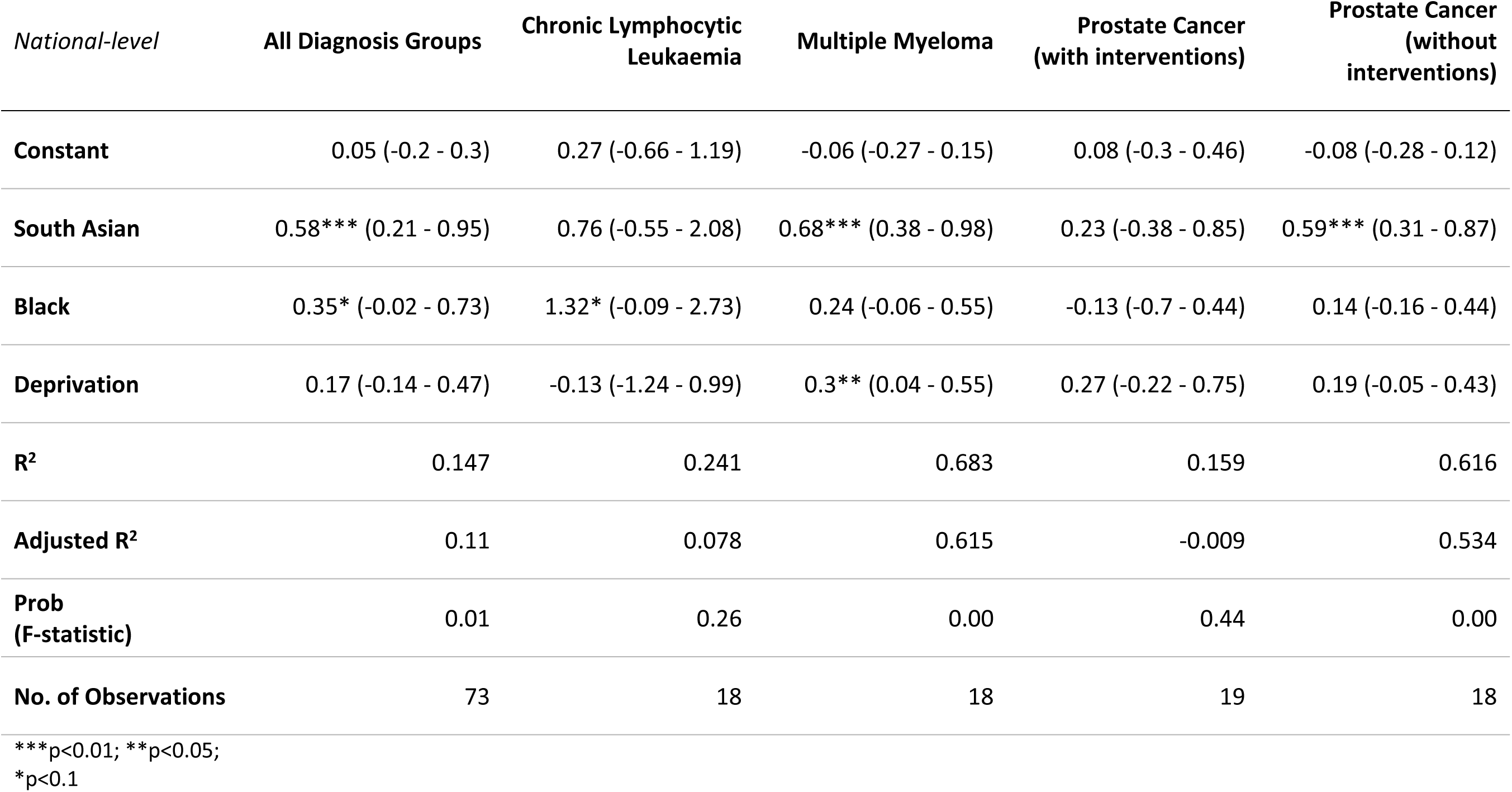
National differential in average normalised CC score (clinical severity) across major cancer types in England.

Table 7B expands the model to look at ICS-level data with additional demographic variables included. Both the South Asian (+0.57, p<0.01) and deprivation (+0.27, p<0.01) binary variables remained statistically significant with the coefficient remaining roughly inline with that observed at a national level. These results are remarkably consistent across different cancer types. Even after controlling for higher levels of deprivation, being of South Asian ethnicity is associated with higher average clinical severity for all but one of the cancers considered in this study. The magnitude of the effect is strongest for more severe forms of prostate cancer (+1.97, p<0.05).

**Table 7B:**
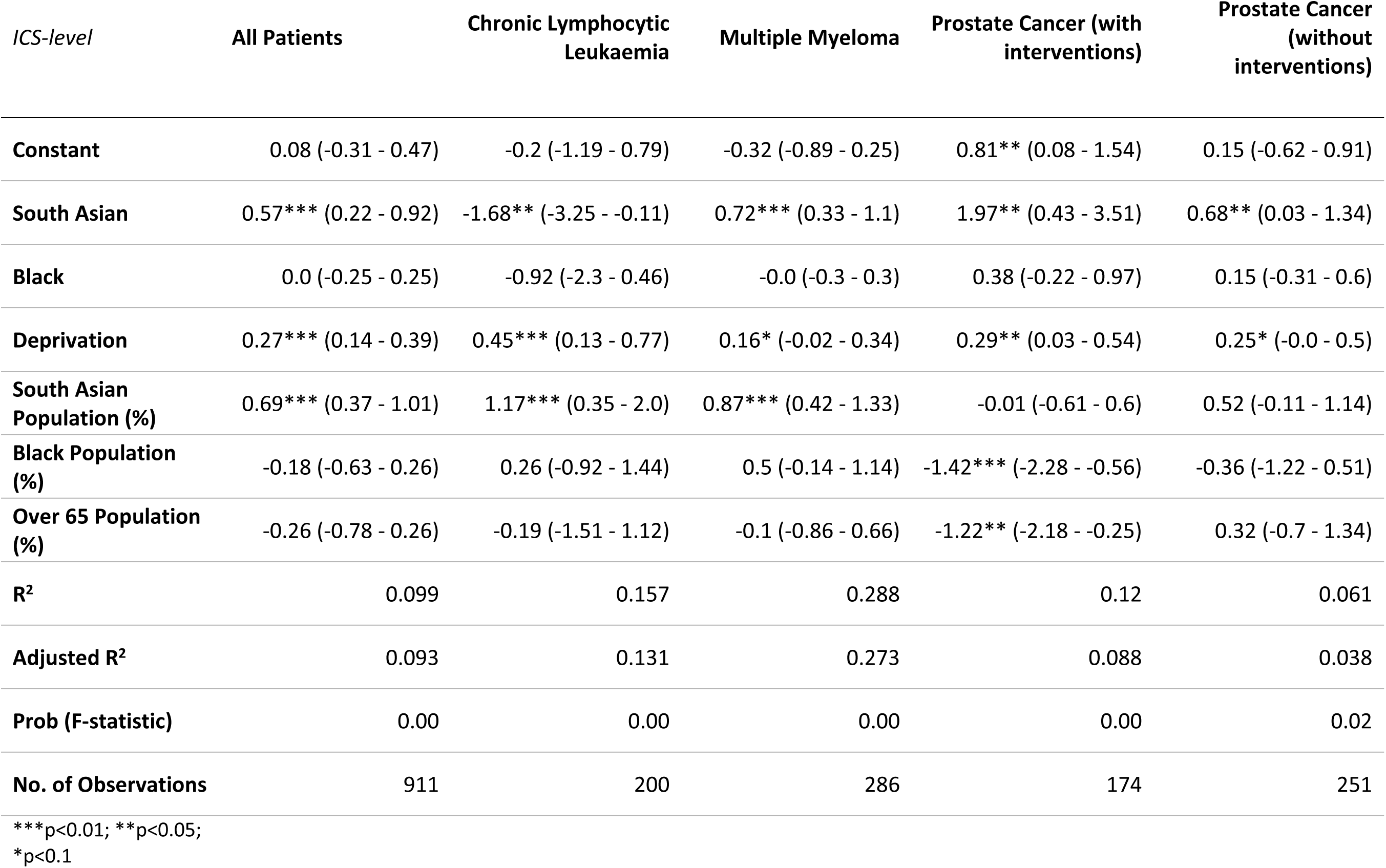
Differential in average normalised CC score (clinical severity) across major cancer types at an Integrated Care System-level in England.

A larger South Asian population, as measured by the percentage of the ICS population of South Asian ethnicity, was associated with higher average clinical severity (+0.69, p<0.01) across all patients, although the effect was only statistically significant for CLL and multiple myeloma. There are several potential drivers for this association including socio-cultural factors specific to South Asian communities. It is also plausible that ICS areas with large South Asian populations share certain socio-economic characteristics associated with greater levels of ill-health and comorbidities.

Black patients do not appear to have statistically higher clinical severity (when controlling for deprivation). Indeed, a higher Black population percentage was associated with lower average clinical severity for prostate cancer with interventions (-1.42, p<0.01). This was also the case in areas with larger elderly populations (-1.22, p<0.05).

### Cost per Patient

Table 8 shows average treatment costs by ethnicity. Treatment costs for black patients are between 23.9 percent and 32.8 percent higher on a national level than the average cost per patient. Average cost per patient are also higher for South Asian (+9.3 percent) and White (Other) patients (+11.5 percent). There is a the strong linear relationship between CC scores and treatment costs. This is to be expected given the role of CC scores in the NHS payments system. However, given that South Asian patients typically have more severe disease, the differential in treatment costs requires further explanation.

**Table 8:**
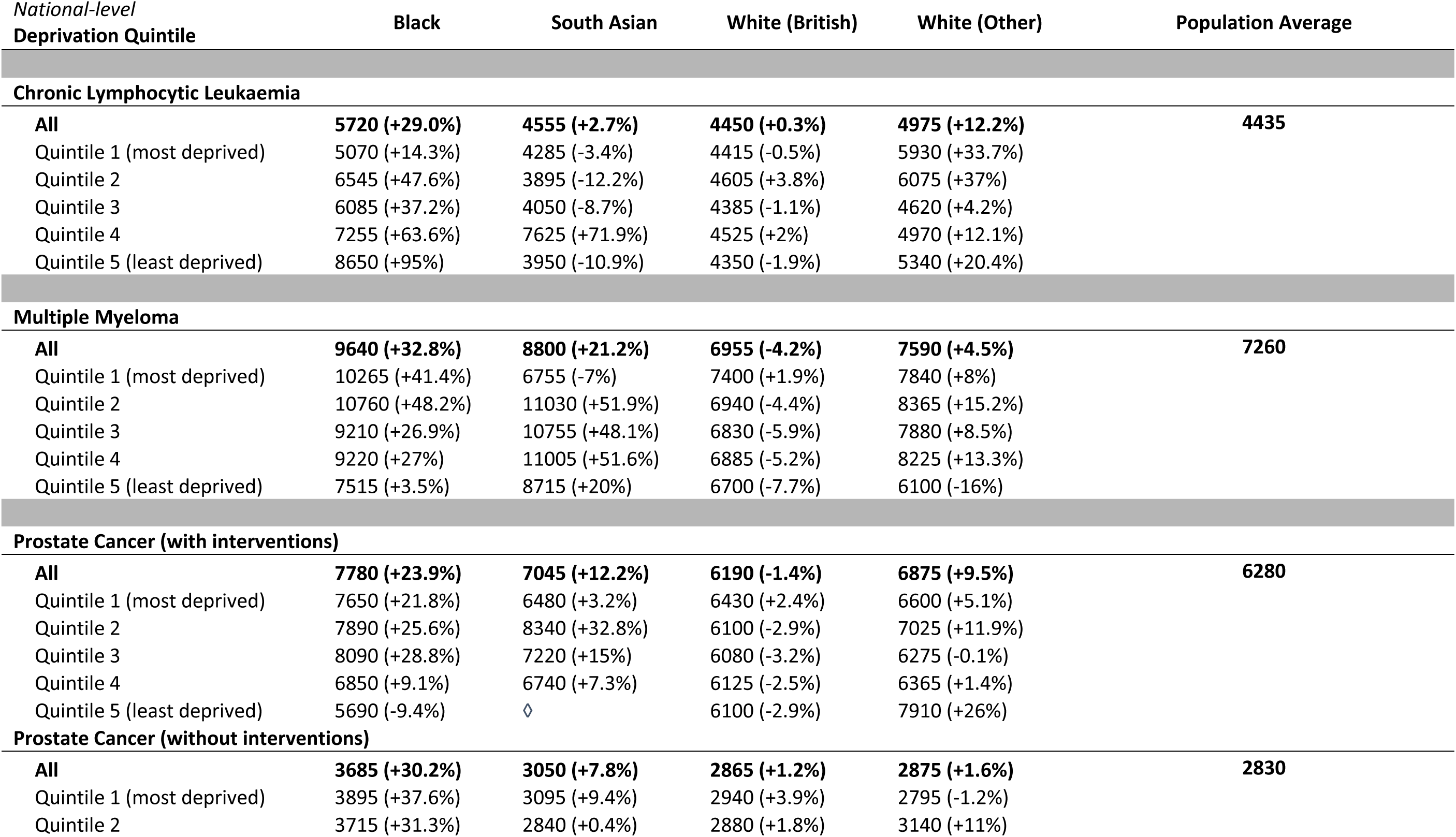
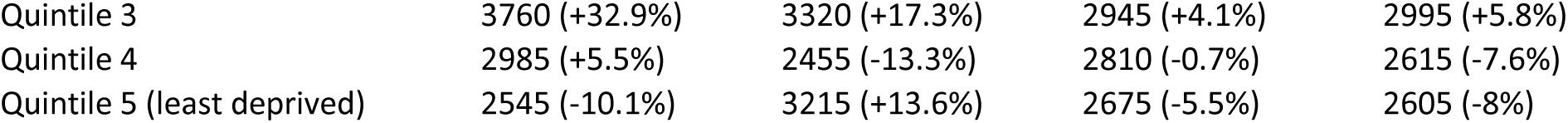
National-level average cost per patient in GBP by ethnicity and deprivation quintile and percentage differential with population average by major ethnic group in England.

Table 9 presents the results of an ICS-level model of the differential in cost per patient from the population average. In addition to the ethnicity, deprivation and the percentage of the population aged over 65 variables described above, a London binary variable was also included. The Black population in England is highly concentrated in a relatively small number of ICS areas, over half the population live in Greater London. The NHS payments system takes account of the higher cost of delivering services in the capital. The London binary seeks to control for the impact of these higher costs.

**Table 9:**
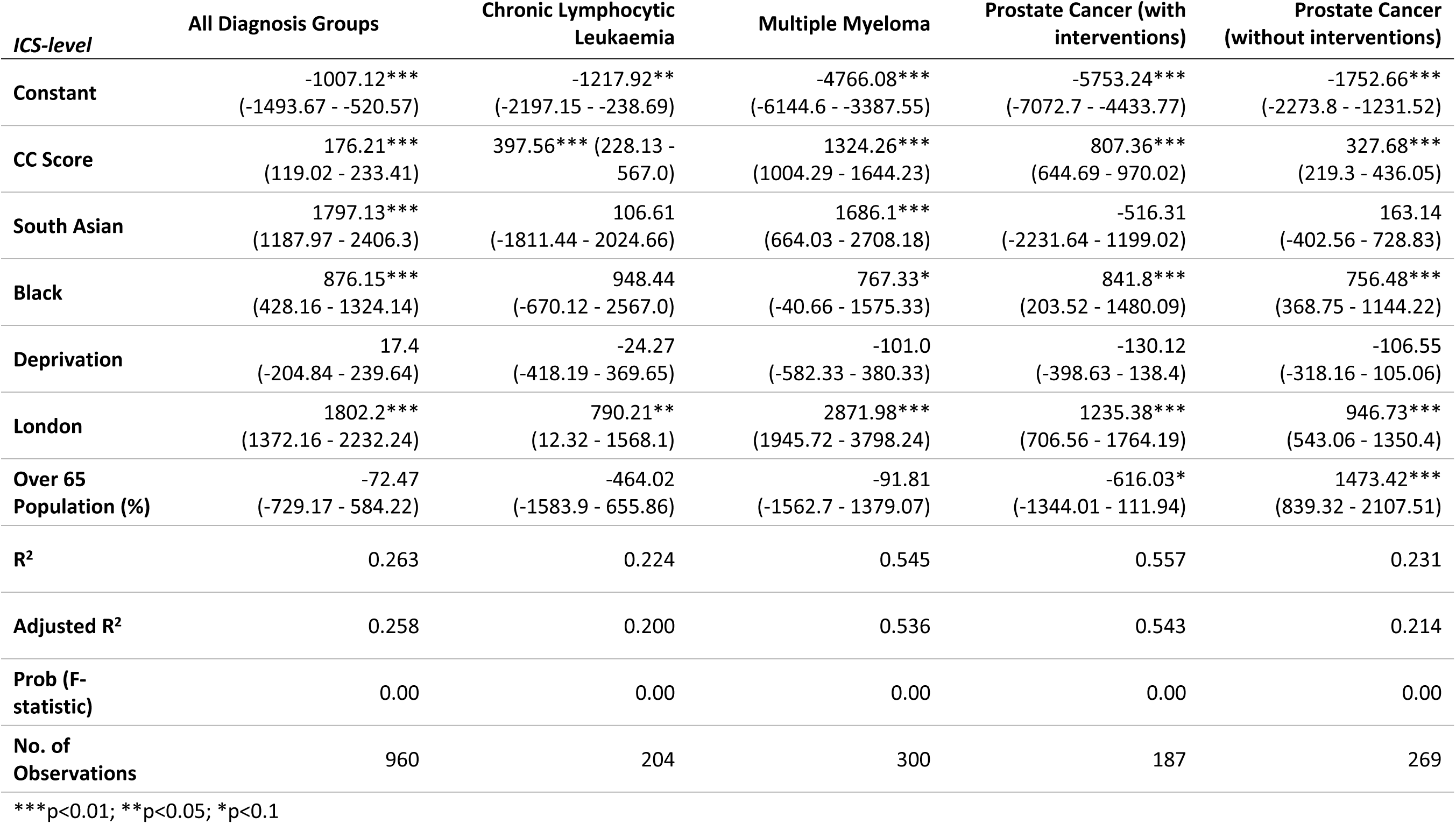
Differential in average cost per patient across major cancer types at an ICS-level in England.

The combined all patient model has reasonably strong explanatory power (R^2^ = 0.24) and accounts for more than half the variation in the disease area specific models for multiple myeloma and prostate cancer with interventions. As expected higher average CC scores and being located in London are statistically significant for all disease areas. Having a larger share of the population aged over 65 was associated with lower average costs, although this was not statistically significant.

After controlling for these variables, being of South Asian (+1797, p<0.01) or Black (+876, p<0.05) ethnicity was associated with higher average costs per patient, although the association was somewhat mixed for South Asian patients. The variable was only statistically significant for multiple myeloma (+1686, p<0.01), although in this case the magnitude of the effect was reasonably large. In contrast, there appears to be a consistent relationship between Black patients and higher treatment costs. Ceteris paribus, treatment costs for Black prostate cancer patients (with interventions) are £842 greater than the national average.

### Patient Activity Indicators

Three patient activity indicators were considered – the number of spells in hospital, mean length of stay and average wait duration. Table 10 displays the average value and percentage difference from the national population average. Black patients appear to have somewhat more spells per patient whereas the number is slightly lower for South Asian patients. All ethnic minority groups appear to have longer periods in hospital than White British patients although, the differential is significantly larger for Black patients. Average wait durations exhibit a high-degree of variation across ethnic groups and cancer types but without a consistent trend. Ethnic minority prostate cancer patients have shorter waits for in-patient appointments, but this pattern is reversed for multiple myeloma patients.

**Table 10:**
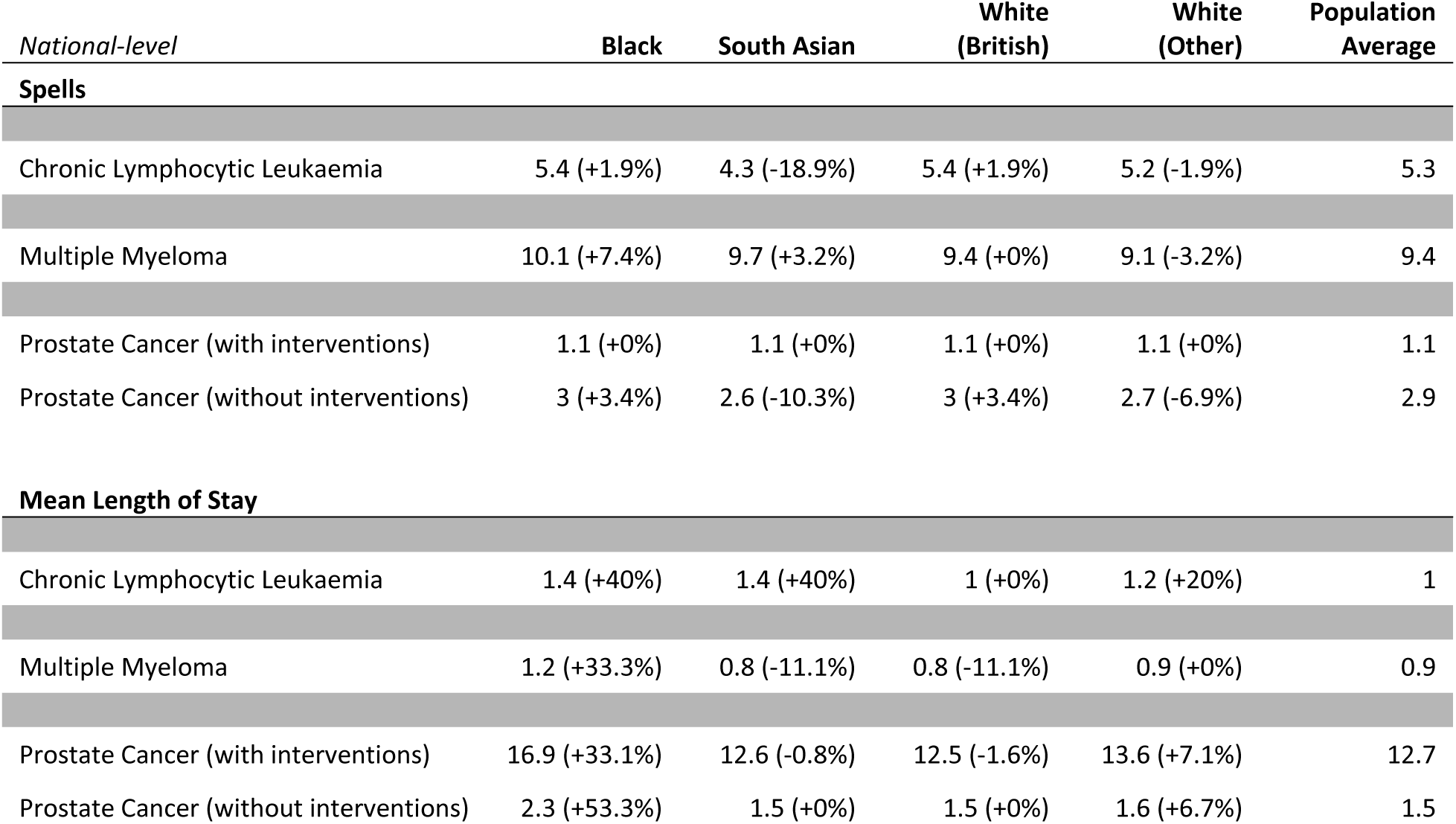
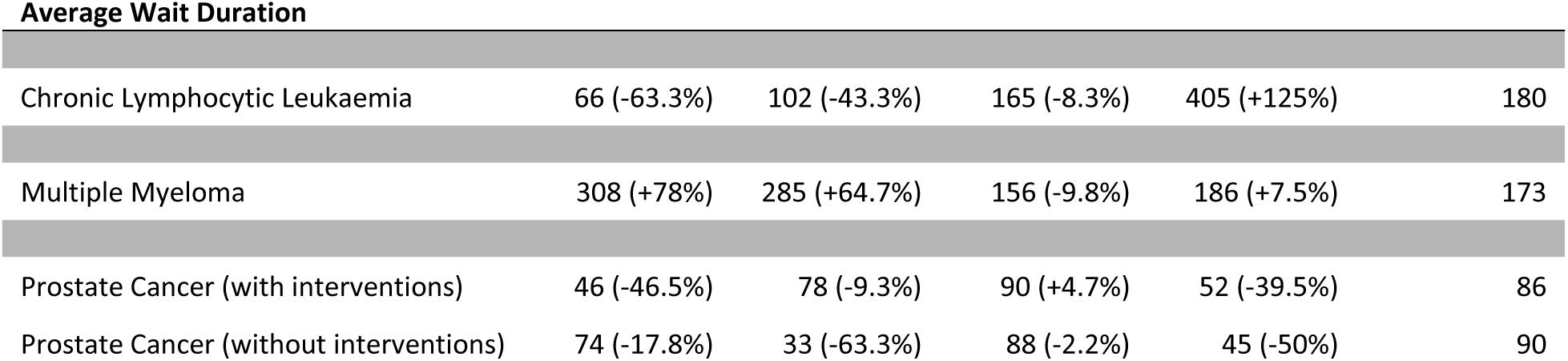
National average spells per patient, mean length of stay and average wait duration by cancer type and ethnicity.

Table 11A and Table 11B presents the results of ICS-level regressions for mean length of stay and spells per patient. Due to the variation in treatment processes across disease areas, it is not appropriate to produce an ‘all patient’ model for patient activity indicators. Results for the average wait duration are not presented as the model was not statistically significant for any of the disease areas (see Table S1). This null result is interesting as it suggests that once a patient has been diagnosed and entered the secondary care system the length of time a patient waits for an appointment is not related to their ethnicity or socio-economic background.

**Table 11A:**
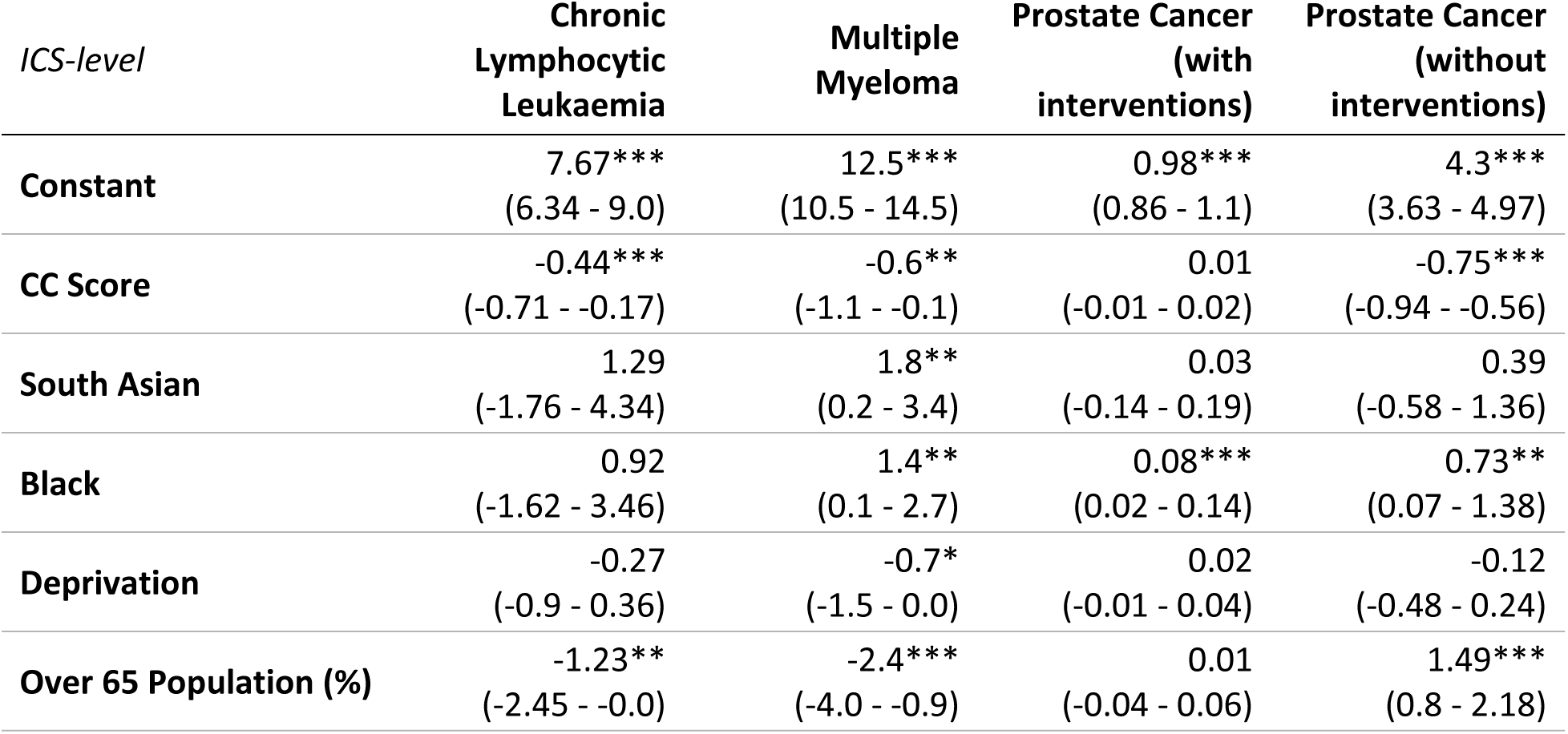
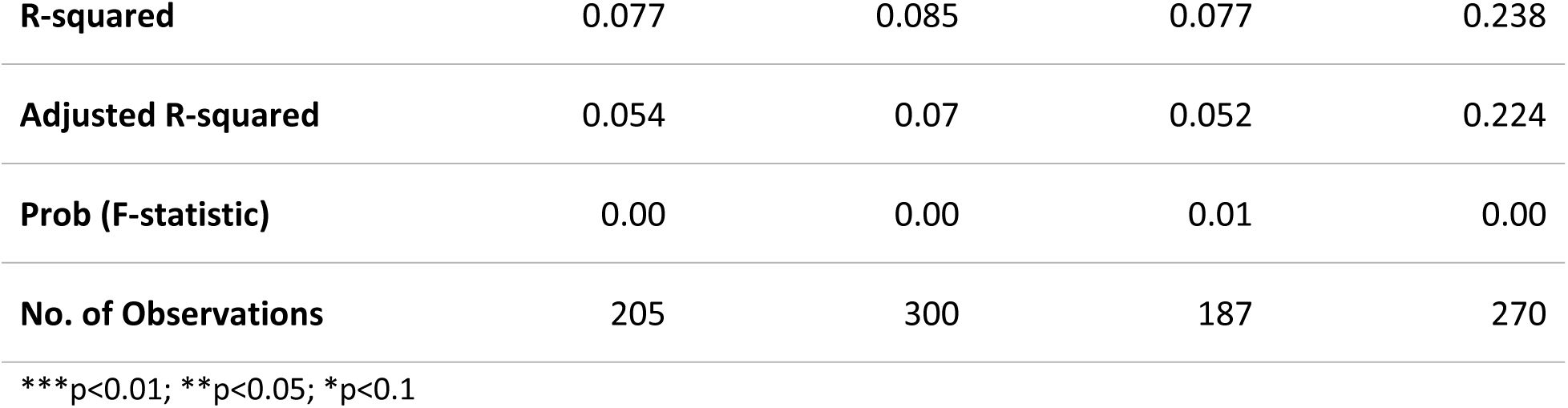
Differential in spells per patient across major cancer types at an Integrated Care System-level.

**Table 11B:**
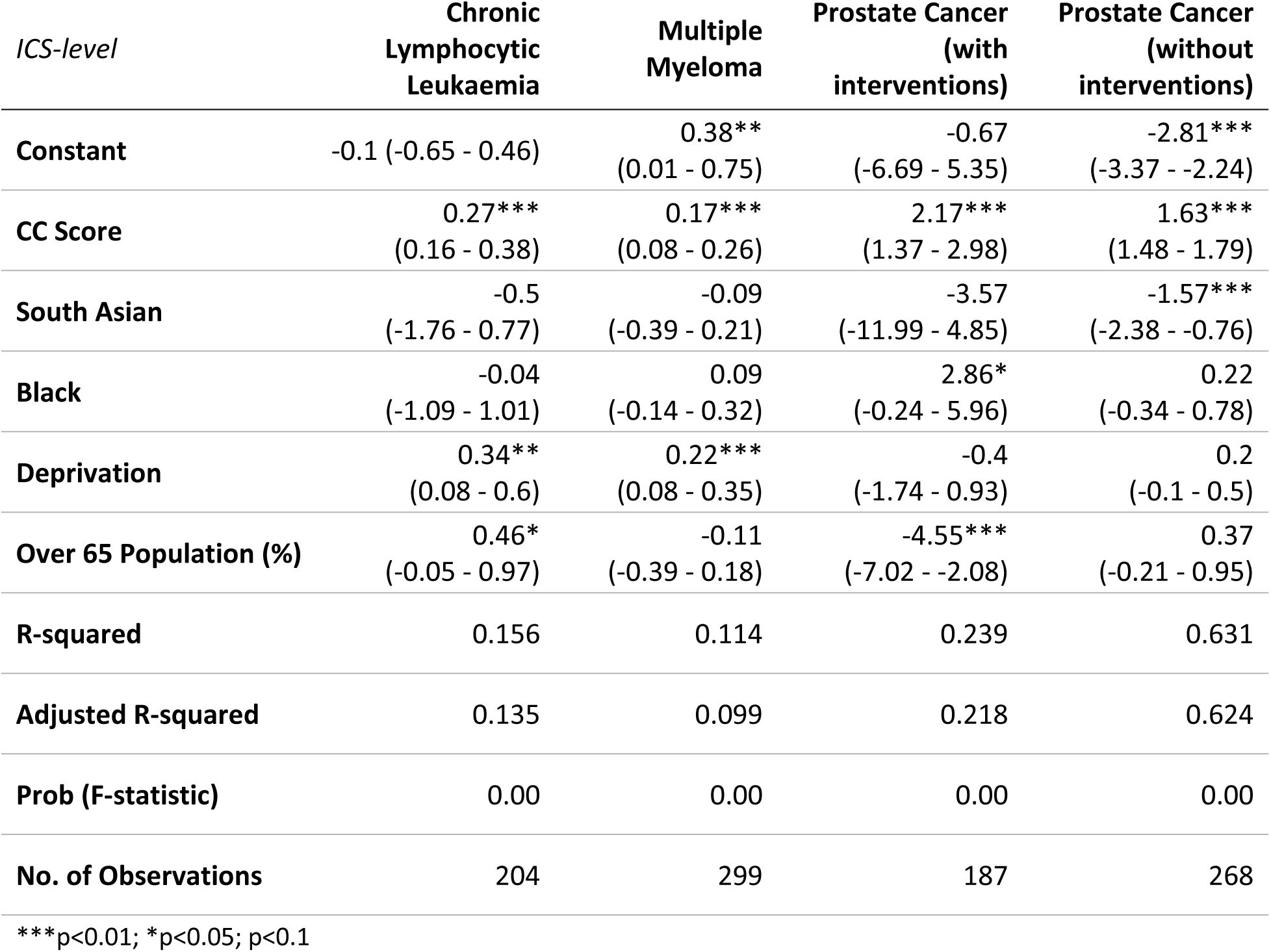
Differential in mean length of stay across major cancer types.

**Table 12:**
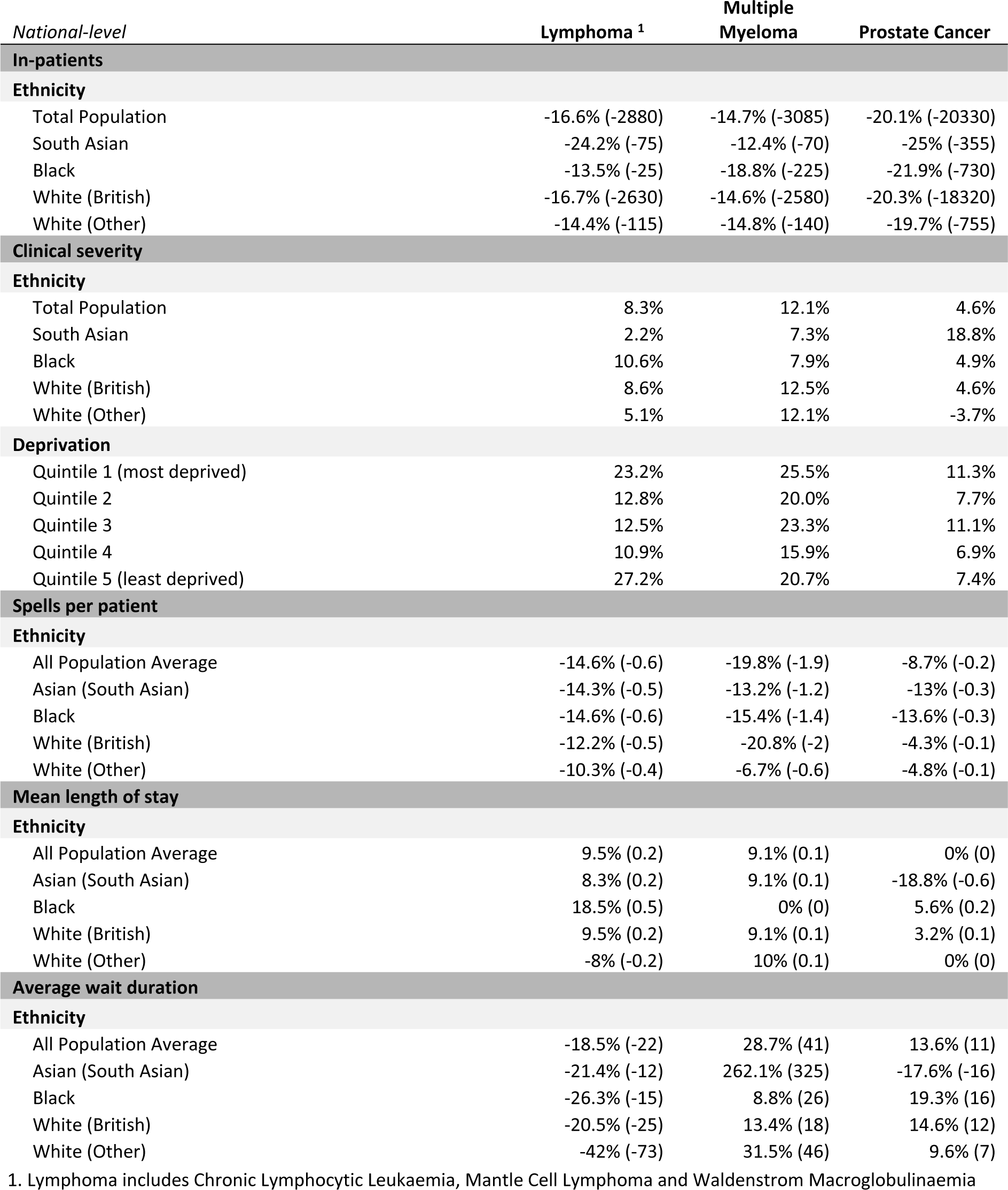
Change in in-patients, clinical severity, spells per patient, mean length of stay and average wait duration between 2019/20 and 2020/21 (COVID-19) in England.

In contrast, both mean length of stay and the number of in-patients spells had a number of variables which met statistical significance. Higher CC scores were associated fewer spells in hospital but a greater average length of stay. This could be associated with more advanced stage of disease amongst populations with higher clinical severity. The relationship is more mixed for ICS areas with older populations although a number of the associations are statistically significant.

Both Black and South Asian patients tend to have a greater number of spells in hospital. The result is reasonably consistent for black patients, however, it is only statistically significant for South Asian multiple myeloma patients (+1.8, p<0.05). Neither ethnic group displayed a consistent pattern for average length of stay. South Asian prostate cancer appear to have shorter average stays, where as Black patients have longer periods in hospital.

### The impact of COVID-19 on ethnic minority cancer patients

Covid-19 placed enormous strain on the NHS and resulted in significant widespread cancellations of non-urgent procedures. Waiting lists and waiting times have increased dramatically. The number of people waiting for elective care in England rose from 4.4 million before the start of the pandemic to over 6 million in February 2022 [37]. The data used in this paper covers the period between the fiscal years of 2016/17 and 2020/21. Consequently, it provides a unique opportunity to analyse the impact of COVID-19 on three cancer types. With the exception of data on average clinical severity, an enlarged version of the original dataset was used which covered all patients with a relevant diagnosis during the study period, without filtering for results with CC scores. Note that unfortunately in this enlarged dataset CLL was not broken out independently, instead a broader definition of ‘lymphoma’ was used which included mantle cell lymphoma and Waldenström macroglobulinaemia as well as CLL. CLL is estimated to account for around two-thirds of the patients in this grouping. Despite this, it was decided to include these results as they provide a clearer view of the total fall in patient numbers as a result of COVID-19.

The fiscal year 2020/21 saw the number of total number of patients fall by 18.9 percent year-on-year. The largest drop in patient number, both in absolute and percentage terms was for prostate cancer. There were 20,330 in-patients (-20.1 percent) fewer than the previous year. The smallest percentage fall was for multiple myeloma (-14.7 percent), the differential may be the result of significantly higher rates of emergency presentation amongst those diagnosed with the disease (see Table 2). Table 11 presents a summary of patient activity indicators split by ethnicity at national level. Due to the small size of the patient populations in individual years no explanatory statistical analysis performed.

Across all disease areas, Black and South Asian patients saw a larger percentage fall in patient numbers than the national average. The number of Black and South Asian cancer patients fell by 1.9 and 2.9 percentage points more than the national average respectively. South Asian patients experienced the greatest percentage fall in patient numbers for both lymphoma (-24.2 percent, 7.63 percentage point deviation from national average)and prostate cancer (-25.0 percent, 4.85 percentage point deviation). Black patients saw the largest fall in multiple myeloma patients (-18.8 percent, 4.17 percentage point deviation).

Average clinical severity increased by 14.7 percent year-on-year. The strength of the differential impact of COVID-19 between ethnicity and clinical severity varied across disease area. The largest increase was amongst South Asian prostate cancer patients, +18.8 percent compared to a national average of +4.6 percent. Black patients saw the largest increase in the severity of CLL. However, White British patients experience the greatest increase in multiple myeloma severity. Overall, the ratio between the population average and the average CC scores of Black and South Asian patients stayed broadly comparable to its pre-pandemic level as clinical severity increased for all ethnic groups.

Patients from the two most deprived quintiles saw a proportionately greater increase in clinical severity (+19.9 percent) than patients in the two least deprived quintiles (+13.4 percent). The effect was greatest amongst South Asian patients for whom more deprived patients saw clinical severity increase by 22.7 percent whereas the least deprived saw clinical severity improve by 14.7 percent.

The impact of COVID-19 can be seen on a range of patient activity indicators. The number of spells per patient fell by 14.37 percent. Counterintuitively, the average length of stay increased slightly for most cancer types and ethnic groups, however, this likely reflects the increase in clinical severity, in turn a reflection of the cancellation of less urgent cases. Waiting times for in-patient appointments displayed considerable variation. Waiting times for lymphoma fell by 18.5 percent, however they increased for both multiple myeloma (+28.7 percent) and prostate cancer (+13.6 percent), although South Asian patients saw their waits fall by 17.6 percent possibly reflecting their higher clinical severity.

## Discussion

Confirming previous research, this study clearly shows the continued presence of inequalities in cancer patient outcomes for a range of cancer types. There are three core findings. South Asian patients have significantly higher clinical severity (CC scores) than the population average. Black and South Asian patients have higher average treatment costs, with the effect particularly strong for Black patients. Finally, COVID-19 had a disproportionate impact on ethnic minority patients.

This study is not able to determine the underlying drivers behind the result. However, the lack of a strong statistically significant relationship between ethnicity and the secondary care patient activity indicators suggests that this may reflect barriers to seeking help or accessing primary care services. Delays to diagnosis would be expected to lead to later stage diagnosis resulting in higher average clinical severity and higher treatment costs. Not only does this place a greater disease burden on ethnic minority populations but it also, as suggested by the higher average treatment costs, places a greater resource burden on the NHS.

There is a well established body of research which shows that ethnic minority patients experience greater socio-cultural barriers to accessing cancer care. These include lower awareness of cancer symptoms and warning signs and, heightened physical and emotional barriers such as being too embarrassed, not being confident to talk, and being too busy which cause individuals to delay seeking help [38][39]. Those of South Asian descent had significant higher perceived barriers than other ethnic groups after controlling for age, gender and deprivation. White British reported experienced the lowest barriers [40]. Several studies of cancer screening uptake found lower uptake rates amongst South Asian individuals, especially Pakistani and Bangladeshi women [41][42]. A review of attitudes to cancer amongst South Asian populations in the UK, US and Canada found a range of beliefs including stigma and fatalism reduced help-seeking and the uptake of screening services [43].

Language and issues related to communication are important barriers to accessing cancer care and health services more broadly. It is also key difference between the South Asian and Black populations. Of those aged over 65, 40.5 percent of the South Asian can not speak English well or at all. This rises to as high as 83.5 percent of older Bangladeshi women. In contrast, the equivalent figure for the Black population is just 4.5 percent [44]. Interviews with South Asian women in the North of England regarding breast cancer screening identified not only issues with direct communication and translation, but also issues relating to the comprehension of healthcare information[45].

Similar emotional barriers are observed in a number of studies examining the attitudes of Black men towards prostate cancer and screening. In these studies, prostate cancer was linked to perceptions of masculinity and sexuality, as well as feelings of fear, shame and denial [46][47][48]. However, there does appear to have been some success in increasing screening rates for prostate cancer in Black men which would tend to reduce clinical severity. In a longitudinal study of prostate-specific antigen testing in inner London general practices, Black men were 4 times as likely as White men to be tested after controlling for age, deprivation, BMI and co-morbidities [49].

Despite the broad evidence of socio-cultural barriers, tangible evidence of delays to diagnosis is mixed. Black and Asian patients are significantly more likely than White patients to have three or more pre-referral GP consultations related to the symptoms that were ultimately diagnosed as cancer [28][50]. However, the proportion of cancer’s diagnosed at an early stage (Stage I or II) is higher for Black (55.4 percent) and South Asian (54.8 percent) patients than the national average (52.2 percent) [51]. A 2013 study found some evidence of longer diagnostic intervals for ethnic minority patients, although not for prostate cancer patients [52].

COVID-19 had a significant impact on cancer care for all ethnic groups across all cancer types included in this study. However, there are clearly specific disease areas in which Black and South Asian patients were disproportionately negatively impacted. The sharp increase in clinical severity amongst South Asian prostate cancer patients, coupled with the greatest reduction in patient numbers, suggests the need for immediate attention and the allocation of resources to prevent existing health inequalities worsening over time.

Whilst, it is too early to make definitive statements regarding the long-term impact of COVID-19 on ethnic inequalities in cancer care this study suggests that they are likely to be significant. Black and South Asian cancer patients had significantly higher average clinical severity than White patients prior to the pandemic, that position has been exacerbated by the pandemic. The size of the ‘missing’ patient population in 2020/21 suggests that these issues are likely to persist and potentially worsen over time. Addressing these ethnic inequalities in cancer care will require not only national-level policy focus but tangible local actions that are tailored to the specific needs of ethnic minority populations.

## Conclusion

This study clearly establishes the relationship between clinical severity, treatment costs and ethnicity. After controlling for socio-economic deprivation, South Asian patients have significantly higher clinical severity than other ethnic groups, whilst it costs the NHS significantly more to treat a patient who is Black than a comparable patient who is White. This is important not only because it indicates that these patient populations experience a disproportionately high burden of disease, but also because it suggests that measures to actively remediate the underlying causes of these effects could improve the efficiency of how NHS resources are allocated. This is particularly important given the context of the recovery from the COVID-19 pandemic and finding that ethnic minority patients were disproportionately negatively impacted.

The UK government’s Life Sciences Vision outlines an ambitious goal of unlocking access to health data to support the research and analysis [15]. As discussed throughout this paper, the relationship between ethnicity and the access to and experience of cancer care is influenced by a variety of overlapping socio-economic, cultural and physiological factors. Whilst many of these factors are systemic, policies which enable local inequalities in access to cancer care to be rapidly identified and targeted measures to be developed are vital if the government is to deliver on its stated goal of reducing health inequalities.

Unfortunately, whilst the underlying data exists in the form of patient records, the existing organisational and technological frameworks are not fit for purpose. The design of this study was based on the availability of data from the Hospital Episode Statistics database which provides information on patient activity in the secondary care sector. However, this only provides a static snapshot at one interface between a patient and the healthcare system. To identify the exact nature and location of the barriers that ethnic minority patients face requires data from each node in the care pathway to be joined together and analysed holistically. This data then needs to be made available in as close to real-time as possible to local NHS commissioning groups to enable them to target resources at the specific needs of local ethnic minority communities.

## Data Availability

The database analysed in this paper was acquired under license from Wilmington Healthcare Ltd. The terms of the license do not permit the reproduction of the database. The database is based on the Hospital Episode Statistics (HES) available from NHS Digital or Wilmington Healthcare. Wilmington Healthcare Ltd. can be contacted by telephone on 0845 121 3686, by e-mail at or by visiting www.wilmingtonhealthcare.com. The data is based on the following ICD-10 code C911 - Lymphoid leukaemia C900 - Multiple myeloma and malignant plasma cell neoplasms Multiple myeloma C61X - Malignant neoplasm of prostate The following HRG Groups: - Chronic Lymphocytic Leukaemia, including Related Disorders - Plasma Cell Disorders - Kidney, Urinary Tract or Prostate Neoplasms, with Interventions - Kidney, Urinary Tract or Prostate Neoplasms, without Interventions Wilmington Healthcare extracted the following aggregated tables for the fiscal years 2020/2021, 2019/2020, 2018/2019, 2017/2018, 2016/2017. Tables: "Population data: Population breakdown by National and ICS level for ethnic groups. ICS Codes & Map: BAME % of Total Population Summary data: National and ICS level inpatient and outpatient activity for patients with a haematology, immunology or oncology diagnosis by fiscal year and for all 5 fiscal years combined Summary data by ethnicity: National and ICS level inpatient and outpatient activity for patients with a haematology, immunology or oncology diagnosis by ethnicity and fiscal year and for all 5 fiscal years combined Summary data by age group: National and ICS level inpatient and outpatient activity for patients with a haematology, immunology or oncology diagnosis by age group for all 5 fiscal years combined Summary data by gender: National and ICS level inpatient and outpatient activity for patients with a haematology, immunology or oncology diagnosis by gender for all 5 fiscal years combined Summary data by IMD: National and ICS level inpatient and outpatient activity for patients with a haematology, immunology or oncology diagnosis by deprivation decile and quintile (using Lower Layer Super Output Area (LSOA) of the patient's residence) for all 5 fiscal years combined Summary data by ethnicity IMD: National level and ICS level inpatient and outpatient activity for patients with a haematology, immunology or oncology diagnosis by ethnicity and deprivation decile (using Lower Layer Super Output Area (LSOA) of the patient's residence) for all 5 fiscal years combined" "HRG data: National and ICS level inpatient activity for patients with a haematology, immunology or oncology diagnosis by year, HRG grouping and average CC score (see HRG Codes tab for details of codes used). The average CC score is calculated from the derived CC score (the mid-point of score ranges or the known value for scores without ranges, e.g. 15+). The average normalised CC score have been normalised on a 1-10 scale. HRG data by ethnicity: National and ICS level inpatient activity for patients with a haematology, immunology or oncology diagnosis by year, ethnicity, HRG grouping, average CC score and average normalised CC score. HRG data by IMD : National and ICS level inpatient activity for patients with a haematology, immunology or oncology diagnosis by year, deprivation quintile (using Lower Layer Super Output Area (LSOA) of the patient's residence), HRG grouping , average CC score and average normalised CC score. HRG data by ethnicity IMD: National and ICS level inpatient activity for patients with a haematology, immunology or oncology diagnosis by year, deprivation quintile, HRG grouping, average CC score and average normalised CC score." "Normalised data: National and ICS level inpatient and outpatient activity (patients, spells, appointments) for patients with a haematology, immunology or oncology diagnosis by year, normalised by 100,000 population using ONS mid-year population estimates Normalised data by ethnicity: National and ICS level inpatient and outpatient activity (patients, spells, appointments) for patients with a haematology, immunology or oncology diagnosis by year and ethnicity, normalised by 100,000 population using ONS mid-year population estimates for ethnicity and organisation Normalised by gender: National and ICS level inpatient and outpatient activity (patients, spells, appointments) for patients with a haematology, immunology or oncology diagnosis by year and gender, normalised by 100,000 population using ONS mid-year population estimates for gender and organisation Normalised data by age group: National and ICS level inpatient and outpatient activity (patients, spells, appointments) for patients with a haematology, immunology or oncology diagnosis by year and age group, normalised by 100,000 population using ONS mid-year population estimates for age group and organisation " "Procedures data: National and ICS level inpatient activity for patients with a haematology, immunology or oncology diagnosis by year and intervention type (see Procedure Codes tab for details of OPCS codes used for each diagnosis group) Procedures data by ethnicity: National and ICS level inpatient activity for patients with a haematology, immunology or oncology diagnosis by year, intervention type and ethnicity Procedures data by IMD: National and ICS level inpatient activity for patients with a haematology, immunology or oncology diagnosis by year, intervention type and deprivation quintile (using Lower Layer Super Output Area (LSOA) of the patient's residence) Procedures data by ethnicity IMD: National and ICS level inpatient activity for patients with a haematology, immunology or oncology diagnosis by year, intervention type, ethnicity and deprivation quintile (using Lower Layer Super Output Area (LSOA) of the patient's residence)" "Covid data: National and ICS level inpatient and outpatient activity for patients with a haematology, immunology or oncology diagnosis for Covid year and 2 year prior average. Absolute and percentage difference between Covid year and 2 year prior average. Covid data by ethnicity: National and ICS level inpatient and outpatient activity for patients with a haematology, immunology or oncology diagnosis and ethnicity for Covid year and 2 year prior average. Absolute and percentage difference between Covid year and 2 year prior average. Covid data by age group: National and ICS level inpatient and outpatient activity for patients with a haematology, immunology or oncology diagnosis and age group for Covid year and 2 year prior average. Absolute and percentage difference between Covid year and 2 year prior average. Covid data by gender: National and ICS level inpatient and outpatient activity for patients with a haematology, immunology or oncology diagnosis and gender for Covid year and 2 year prior average. Absolute and percentage difference between Covid year and 2 year prior average. Covid data by IMD: National and ICS level inpatient and outpatient activity for patients with a haematology, immunology or oncology diagnosis by deprivation decile and quintile (using Lower Layer Super Output Area (LSOA) of the patient's residence) for Covid year and 2 year prior average. Absolute and percentage difference between Covid year and 2 year prior average. Covid data by ethnicity IMD: National and ICS level inpatient and outpatient activity for patients with a haematology, immunology or oncology diagnosis by ethnicity and deprivation decile and quintile (using Lower Layer Super Output Area (LSOA) of the patient's residence) for Covid year and 2 year prior average. Absolute and percentage difference between Covid year and 2 year prior average."

## Appendix

**Table S1.**
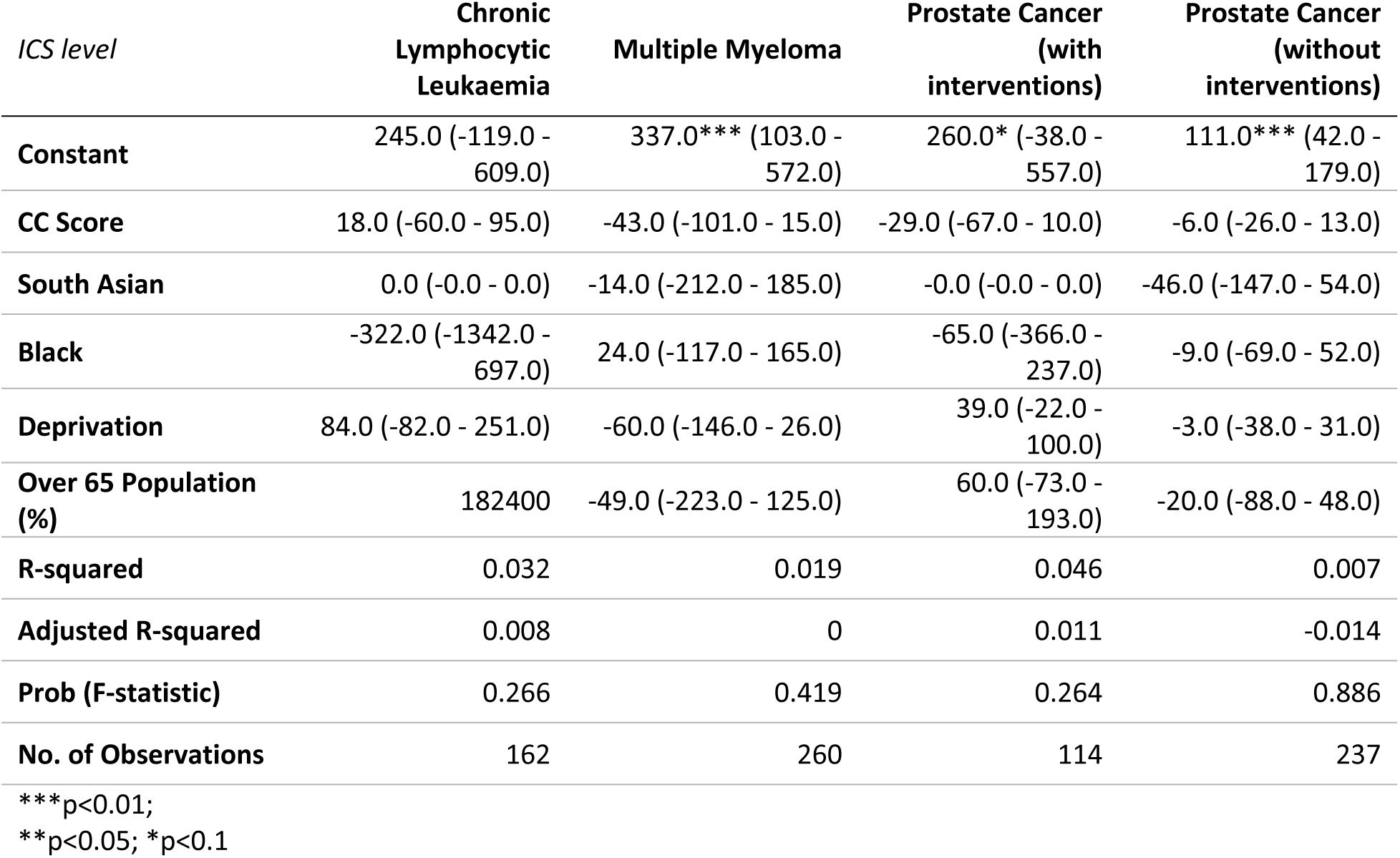
ICS-level differential in average wait duration across major cancer types.

